# Survival analysis of hospital length of stay of novel coronavirus (COVID-19) pneumonia patients in Sichuan, China

**DOI:** 10.1101/2020.04.07.20057299

**Authors:** Zhuo Wang, John S. Ji, Yang Liu, Runyou Liu, Yuxin Zha, Xiaoyu Chang, Lun Zhang, Qian Liu, Yu Zhang, Jing Zeng, Ting Dong, Xinyin Xu, Lijun Zhou, Jun He, Yin Deng, Bo Zhong, Xianping Wu

## Abstract

**Objective:** Allocation of medical resource is essential to a strong public health system in response to COVID-19. Analysis of confirmed COVID-19 patients’ hospital length of stay in Sichuan can be informative to decision-making in other regions of the world.

**Design:** A retrospective cross-sectional study.

**Data and Method:** Data from confirmed COVID-19 cases in Sichuan Province were obtained from the National Notifiable Diseases Reporting System (NNDRS) and field survey. We collected information on demographic, epidemiological, clinical characteristics, and the length of hospital stay for confirmed patients. We conducted an exploratory analysis using adjusted multivariate cox-proportional models.

**Participants:** A total of 538 confirmed patients of COVID-19 infection in Sichuan Province from January to March 2020.

**Outcome measure:** The length of hospital stay after admissions for confirmed patients.

**Results:** From January 16, 2020 to March 4, 2020, 538 human cases of COVID-19 infection were laboratory-confirmed, and were hospitalized for treatment. Among these, 271 (50%) were 45 years of age or above, 285 (53%) were male, 450 (84%) were considered as having mild symptoms. The median hospital length of stay was 19 days (interquartile range (IQR): 14-23, Range: 3-41). Adjusted multivariate analysis showed that longer hospital length of stay was associated with factors aged 45 and over (HR: 0.74, 95% CI: 0.60-0.91), those admitted to provincial hospital (HR: 0.73, 95% CI: 0.54-0.99), and those with serious illness (HR: 0.66, 95% CI: 0.48-0.90); living in areas with more than 5.5 healthcare workers per 1000 population (HR: 1.32, 95% CI: 1.05-1.65) was associated with shorter hospital length of stay. There was no gender difference.

**Conclusions:** Preparation control measures of COVID-19 should involve the allocation of sufficient medical resources, especially in areas with older vulnerable populations and in areas that lack basic medical resources.

**Strengths and limitations of this study:**

▸ Patients at least 45 years, those with serious illness, those living in areas with fewer healthcare workers per 1,000 people, and those admitted to higher levels of hospitalization had longer lengths of hospitalization, while gender, time interval from onset to visit the hospital had no effect on the length of the hospital stay.
▸ Preparation of timely evidence-based prevention and control measures for COVID-19 involve allocation of sufficient medical resources, especially in areas with older vulnerable populations and in areas that lack basic medical resources.
▸ Based on findings, it is of great significance to strengthen the construction of multi-level medical institutions in response to public health emergencies and occupation of medical resources.
▸ The characteristics of inpatients can be further subdivided to obtain more detailed inpatient characteristics.

## INTRODUCTION

Sichuan Province has a population of more than 80 million people. Despite of being located adjacent to Hubei Province, where the COVID-19 transmission originated,^1-3^ the number of confirmed cases of COVID-19 was fewer than 600 as of March, 2020. This is partially due to preventive measures taken by government authorities and residents quarantine measures.

Thus far, many studies have reported COVID-19 epidemiological and clinical features, molecular and biological mechanisms, prevention and control management.^4-6^ These descriptive studies have allowed researchers and policy makers to understand the incubation period and transmutability of SAR-CoV-2^7 8^. As of now, knowledge on factors affecting the hospital length of stay is evolving. As the global pandemic of COVID-19 progress in other regions of the world, insight into hospital length of stay is informative for planning of allocation of medical resources to those most in need.

In this study, the relationship between demographic characteristics of confirmed patients, individual treatment behavior, local medical resources, hospital grade and length of stay in hospital were assessed in relation to hospital length of stay in Sichuan.

## METHODS

### Data sources

We used data on laboratory-confirmed cases of COVID-19, which were reported to the Sichuan Center for Disease Control and Prevention (CDC) through the National Notifiable Diseases Reporting System (NNDRS).^9^ In addition to the network of data reported by hospitals at all levels, part of the data come from a field survey of confirmed cases conducted by the staff of local or provincial CDC and Health Resources Report of Health Commission of Sichuan Province.^10^

Demographic, epidemiological, and basic clinical data were obtained by combining these data sources. Combined the information of 538 confirmed patients in Sichuan Province from January to March 2020 was used for analysis, which included age, sex, place of residence, dates of illness onset, dates of diagnosis, hospital admission, discharge, clinical grade, hospital level and health service personnel per 1,000 population. The period of data spanned the first confirmed case in Sichuan Province on January 16, 2020 to March 4, 2020.

### Definitions

Diagnostic and treatment protocol for Novel Coronavirus Pneumonia (Trial version 7) was released by National Health Commission of the People’s Republic of China, which specifies case definitions, diagnosis, differential diagnosis, treatment, laboratory assays and discharge criteria.^11^ A confirmed case was defined by epidemiological history and/or clinical features that a suspected case may contact with COVID-19 infected persons and/or had symptoms and signs of COVID-19 infection, plus positive laboratory test including virus nucleic acid, gene sequencing and serum antibody detection. Clinical grades were divided into four levels: mild, common, heavy and severe, which we combined mild and common as mild illness and heavy and severe as severe illness. The mild or common type was characterized by mild symptoms, with or without fever, respiratory tract symptoms, and image manifestations of pneumonia. Heavy or severe cases were shortness of breath or low oxygen saturation, or even respiratory failure. Discharge criteria were: a. the body temperature returned to normal for more than 3 days; b. the respiratory symptoms improved significantly; c. the pulmonary imaging showed a significant improvement in acute exudative lesions; d. the nucleic acid tests were negative for two consecutive respiratory specimens (sampling interval at least 1 day). Hospital grades were divided into three categories: provincial level, city level and county level, which were managed by different levels of government, and we combine the city and county level into non provincial level.

### Statistical analysis

The relationship between patient’s age, gender, time interval from illness onset to diagnosis, hospital grade of patients, health servicers per 1,000 population in the patient’s permanent residence, clinical grade, and the length of hospital stay after admissions for confirmed patients was compared and analyzed by survival analysis. The event of interest is discharge status, 0 for no discharge, 1 for discharge, and death is treated as 0. The Kaplan-Meier method was used for single factor comparison, and the Cox Proportional Hazards Model was used for multi-factor comparison.^12^ Its formula was:

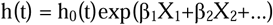

Where h(t) is the discharge hazard at stay length t for a patient with a set of predictors X_1_, X_2_…; h_0_(t) is the baseline discharge hazard function; covariates X were the patient’s age, gender, time interval from illness onset to diagnosis, hospital grade of patients, health servicers per 1,000 population in the patient’s permanent residence, clinical grade, and β_1_, β_2_… are the model parameters describing the effect of the predictors on the overall hazard. We transformed continuous (age, time interval from illness onset to diagnosis, health servicers per 1,000 population) and multilevel variables (hospital grade of patients, clinical grade) into binary variables according to the variables’ distribution. Statistical analyses were conducted in R (version 3.6.3), and p-values less than 0.05 for parameter estimates were considered statistically significant.

### Patient and public involvement

There was no patient or public involvement in the design or data analysis of this study.

## RESULTS

The first confirmed case in Sichuan Province was hospitalized on January 16, 2020. By March 4, 2020, 538 laboratory-confirmed human cases of COVID-19 infection have been hospitalized for treatment in Sichuan Province. Of those, 364 patients recovered and was discharged. Only three patients died. 271 (50%) of the 538 individuals who had to be admitted to hospital were aged at least 45 years, and 285 (53%) were male, 450 (84%) were classified as having mild illness. Among the discharged patients, 150 (41%) took at least 5 days from illness onset to diagnosis, 301 (83%) were admitted to hospitals below the provincial level, and 190 (52%) lived in areas with a medical resource density of more than 5.5 health service personnel per 1000 residents (Table 1).

**Table 1:** Characteristics and median time to stay of 538 COVID-19 confirmed patients who were admitted to hospital

| Group | Discharged<sup>a</sup> | Unresolved/<br>Died | Total | Median estimate of<br>stay length<sup>b</sup> | 95% Confidence interval |  | IQR<sup>c</sup> | Range of stay length |
| --- | --- | --- | --- | --- | --- | --- | --- | --- |
|  |  |  |  |  | Lower<br>limit | Upper limit |  |  |
| Total | 364 | 174 | 538 | 19 | 18 | 21 | 14-23 | 3-41 |
| Age (years) |  |  |  |  |  |  |  |  |
| <45 | 200 (55%) | 67 (39%) | 267 (50%) | 18 | 17 | 19 | 13-22 | 3-41 |
| ≥45 | 164 (45%) | 107 (61%) | 271 (50%) | 21 | 19 | 23 | 14-24 | 3-41 |
| Gender |  |  |  |  |  |  |  |  |
| Male | 192 (53%) | 93 (53%) | 285 (53%) | 19 | 18 | 21 | 14-24 | 3-41 |
| Female | 172 (47%) | 81 (47%) | 253 (47%) | 19 | 18 | 21 | 14-23 | 3-37 |
| Time interval from illness onset to diagnosis (days) |  |  |  |  |  |  |  |  |
| <5 | 214 (59%) | 105 (60%) | 319 (59%) | 19 | 19 | 21 | 14-23 | 3-39 |
| ≥5 | 150 (41%) | 69 (40%) | 219 (47%) | 18 | 17 | 21 | 13-24 | 3-41 |
| Hospital grade |  |  |  |  |  |  |  |  |
| Provincial | 63 (17%) | 56 (32%) | 119 (22%) | 20 | 15 | NA | 10-24 | 3-41 |
| Non-provincial | 301 (83%) | 118 (68%) | 419 (78%) | 19 | 18 | 20 | 14-23 | 3-41 |
| Health servicers per 1,000 population |  |  |  |  |  |  |  |  |
| <5.5 | 174 (48%) | 96 (55%) | 270 (50%) | 20 | 19 | 22 | 15-23 | 5-39 |
| ≥5.5 | 190 (52%) | 78 (45%) | 268 (50%) | 18 | 17 | 21 | 12-23 | 3-41 |
| Clinical grade |  |  |  |  |  |  |  |  |
| Mild illness | 315 (87%) | 135 (78%) | 450 (84%) | 19 | 17 | 19 | 14-23 | 3-39 |
| Severe illness | 49 (13%) | 39 (22%) | 88 (16%) | 23 | 21 | 31 | 15-26 | 3-41 |

a Data are n (%); b The estimate was unadjusted; c IQR: interquartile range.

The median length of stay for all confirmed inpatients was 19 days (interquartile range (IQR): 14-23, range: 3-41), while it was 21 days (IQR: 14-24, range: 3-41) for those aged 45 years and above, 18 days (IQR: 13-22, range: 3-41) for people under 45 years old, 19 days (IQR: 14-24, range: 3-41 for male; IQR: 14-23, range: 3-37 for female) for both men and women, 18 days (IQR: 13-24, range: 3-41) for patients with at least a 5-day interval from illness onset to visit hospital, 19 days (IQR: 14-23, range: 3-41) for patients who were admitted to hospitals below the provincial level, 18 days (IQR: 12-23, range: 3-41) for patients living in areas with a medical resource density greater than 5.5 healthcare workers per 1000 population, and 19 days (IQR: 14-23, range: 3-39) for mild cases (Table 1 and Figure 1A).

**Figure 1:**
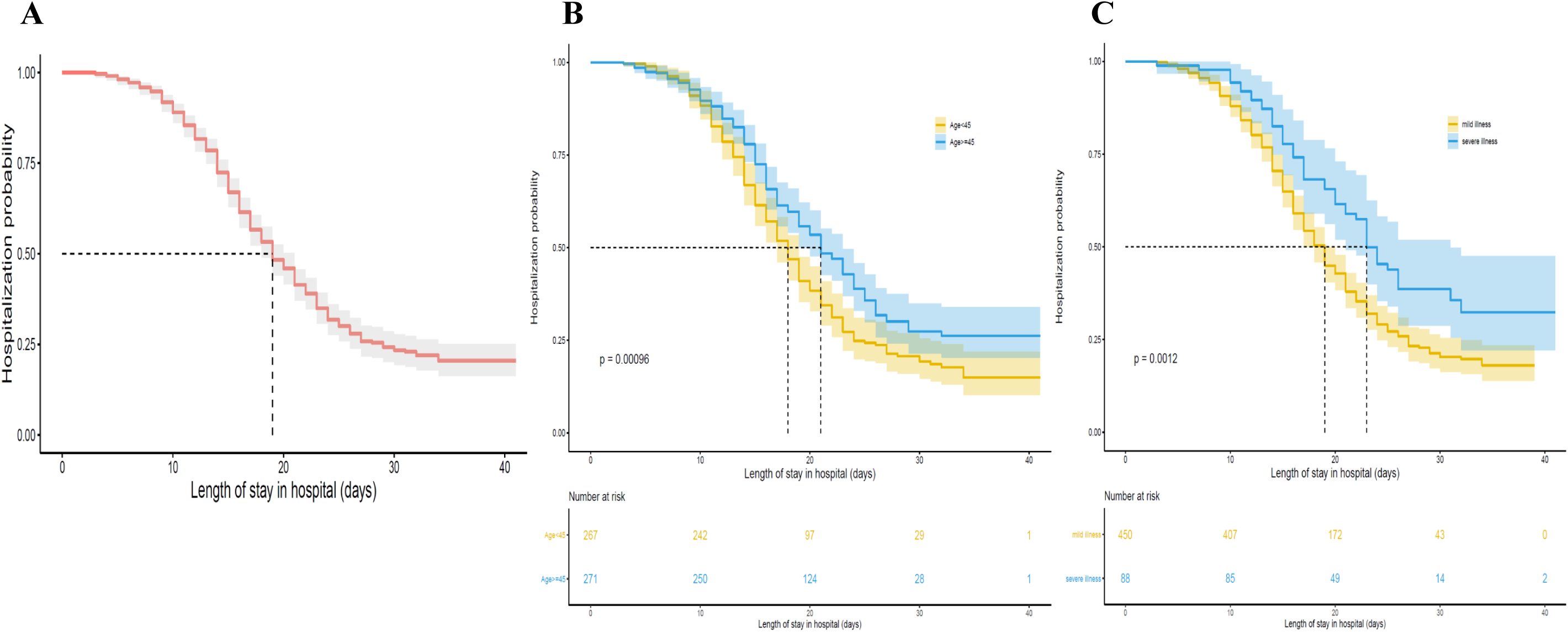
Probability of stay in hospital for COVID-19 confirmed patients. Shading shows 95% CIs. (A) 538 COVID-19 confirmed patients of all ages, (B) 267 patients younger than 45 years and 271 patients at least 45 years, (C) 450 mild cases and 88 severe cases.

We used univariate Kaplan-Meier comparisons and found age and clinical grade were strongly related to stay length (P<0.01, Figure 1B and Figure 1C). Severe patients aged 45 years and over had longer hospital stays. The results of multivariate Cox Proportional Hazards Model showed that being aged 45 years and over (Hazard ratio (HR): 0.74, 95% confidence interval (CI): 0.60-0.91, P= 0.005), being admitted to provincial hospital (HR: 0.73, 95% CI: 0.54-0.99, P= 0.04), living in areas with more than 5.5 healthcare workers per 1000 population (HR: 1.32, 95% CI: 1.05-1.65, P= 0.016), and having serious illness (HR: 0.66, 95% CI: 0.48-0.90, P= 0.008) were the risk factors for prolonged hospital stays. The HR less than one indicates that the probability of discharge was reduced and the risk of prolonged hospitalization was increased (Figure 2). Alternatively, we also considered age and service density as continuous variables and tried a stepwise-selected Cox model to calculate HR. The relationships between these four variables and discharge were still statistically different.

**Figure 2.**
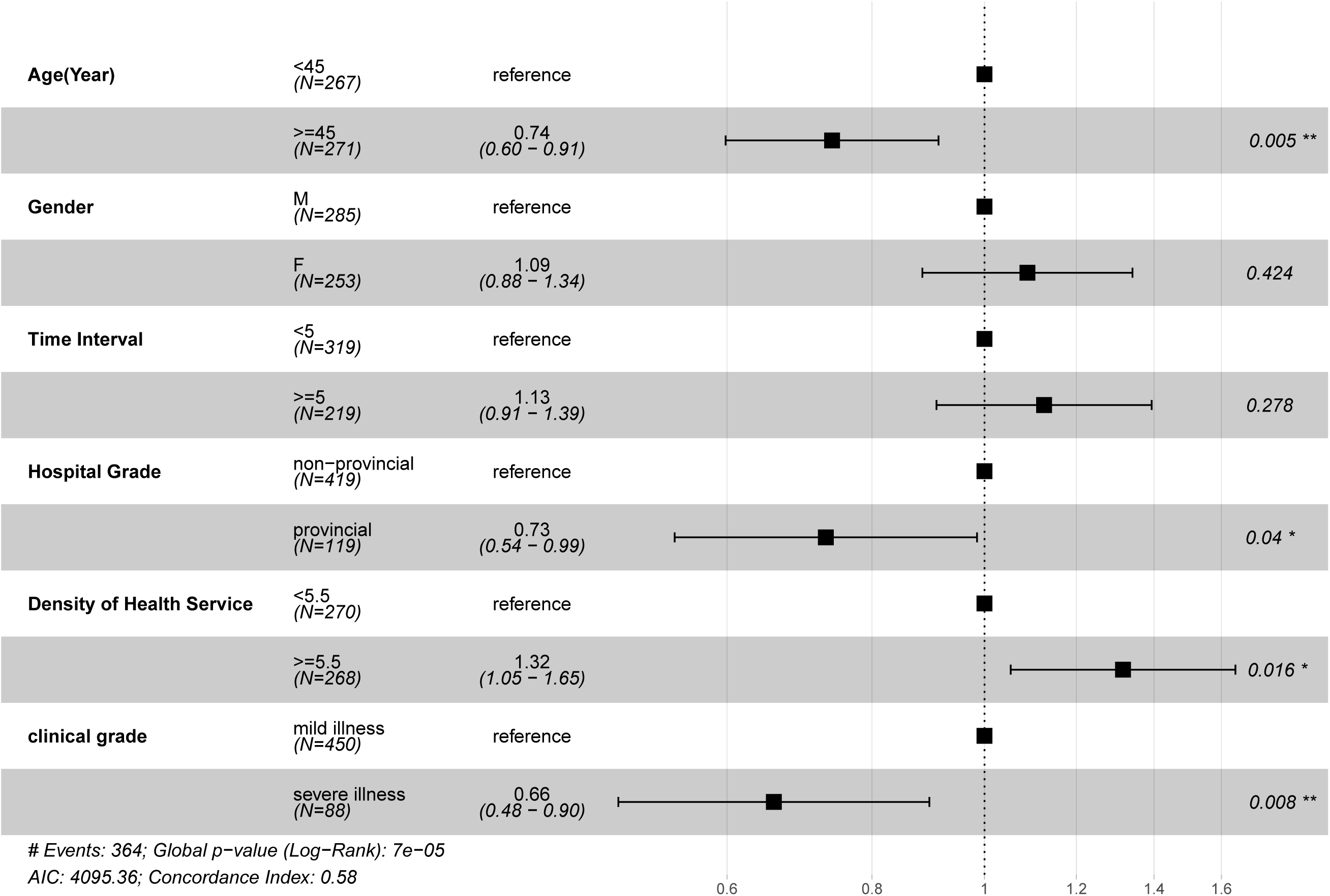
Hazard ratio (HR) of discharge for COVID-19 confirmed patients. Adjusted model includes age, gender, time interval, hospital grade, density of health service and clinical grade.

## DISCUSSION

On December 31, 2019, China reported for the first time to the WHO Country Office an unexplained pneumonia found in Wuhan. On January 30, 2020, the COVID-19 outbreak was announced as a public health emergency of international concern. On March 11, 2020, WHO characterized COVID-19 as pandemic.^13 14^ Novel coronavirus pneumonia has become a major public health event because of its extremely rapid transmission speed, large-scale spread and the extreme utilization of medical resources by numerous patients. Therefore, according to the local demographics and the current status of medical resources, it is extremely important to prepare for risk prevention and control in advance. We modeled demographic characteristics, treatment behaviors, clinical characteristics and local medical resources of hospitalized confirmed patients in order to inform prospective risk assessment for on different areas. Based on the hospitalization data of confirmed patients in Sichuan Province, we found that patients at least 45 years, those with serious illness, those living in areas with fewer healthcare workers per 1,000 people, and those admitted to higher levels of hospitalization had longer lengths of hospitalization, while gender, time interval from onset to visit the hospital had no effect on the length of the hospital stay. That is to say, areas with more vulnerable older populations may require more medical resource allocations to cope with longer hospital stays. In contrast, whether patients see a doctor in time does not seem to affect hospitalization periods, as the results of single factor and multi-factor survival analysis showed that there was no significant difference in the length of stay between the two groups whose time interval from onset to diagnosis is within 5 days and more than 5 day.

Sichuan Province, as a large populous and frequently travelled to region close to Hubei, has some control measures worth sharing with other regions.^15^ On January 21, 2020, the Sichuan provincial government began to take a series of steps towards epidemic prevention and control, including conducting epidemic prevention training for primary healthcare workers, formulating new plans to allocate resources, preparing medical resources that would be used for at least until May, requiring patients with mild illness or suspected patients to select hospitals as close as possible to avoid cross infection, etc.^16^ After the completion of these steps, the “first level emergency response” of the whole province was launched on January 24, 2020, requiring prohibition of any form of group gathering activities and avoidance of public panic.^17^ This series of preparations may also be one of the reasons why the confirmed patients’ median length of stay (19 days) in hospital in Sichuan Province is longer than the national average, which is 10 days, so that patients can get longer treatment and care in the hospital.^1^

In China, the majority of cases were classified as mild (81%). The overall mortality rate was estimated to be 2.3%.^18 19^ Although the mortality rate of COVID-19 was lower than that of SARS (10%) and MERS (34%), the total number deaths of COVID-19 globally exceeded SARS and MERS.^20-22^ We posit that insufficient medical resources or unreasonable allocation of medical resources may be one of the reasons for the excessive number of deaths. For example, many patients with mild illness may occupy many medical resources. In Sichuan Province, 78% (419 / 538) of the patients were admitted to non-provincial hospitals. Single factor analysis showed that different levels of hospitals had no effect on the length of inpatient treatment. After controlling the covariates such as patient age, clinical severity and regional medical service density, we found that the hospital level had a weak association to the length of stay, which was mainly attributed to the clinical severity of patients. And patients in areas with more than 5.5 health servicers per 1,000 population had shorter hospital stays. Massonnaud et al. assessed COVID-19 impact on healthcare resources for each French metropolitan region based on the average length of stay in hospitals of 15 days, and showed that even in the best case scenario, the French healthcare system will very soon be overwhelmed^23^ This paper shows the necessity of establishing a multi-level medical system and strengthening primary medical care, which can reduce the pressure of high-level hospitals in case of public health emergencies. In recent years, China has been strengthening the construction of lower-level medical systems and the monitoring of major diseases and health hazards, which deserve further implementation.^24^

In addition, this study pointed out that the interval length of time from symptoms to doctor’s visit did not affect the length of stay in hospitals, while some patients may mistakenly regard the COVID-19 infection as influenza, but COVID-19 is more infectious and pathogenic,^25^ and the length of stay in hospital is longer, while the mean length of stay for the treatment of influenza was less than a week.^26^ We suggest that in areas with insufficient medical resources and shortage of inpatient beds, patients under 45 years old with mild illness could choose to be treated at home with quarantine under the guidance of doctors, which may avoid cross infection and crowding of medical resources, under the circumstances of access to medical security at home, and timely communication between doctors and patients. While promoting quarantine, best-practices should be considered, such as placing patients in an isolated single room with good ventilation and away from visitors.^27-29^

### Strengths and limitations

As far as we know, this is the first study to report factors affecting the length of stay of COVID-19 from a province in China. We used the original data in Sichuan Province, a place with a size that is comparable to Germany, and more than that of most European countries such as Britain and Italy in population, and per capita gross domestic product is less than 50 thousand yuan, lower than that of most European countries ^30^. According to the research results, our study is a novel finding to make recommendations for policies, measures and strategies.^31^ We believe that the results of this study can serve as a practical reference for global control of COVID-19 under situations of a global pandemic and shortages of medical resources.

There are some limitations of the study. First, there may be other factors that affect the length of stay that are not included in the analysis, such as whether the diagnosed patient has underlying diseases. Second, this study did not further divide the confirmed cases of COVID-19 into imported or local cases, in order to obtain more detailed inpatient characteristics.

## CONCLUSIONS

First, in regions where there are more vulnerable older populations, and where basic medical resources are limited, the government has to make advanced prevention and control measures for COVID-19 to provide sufficient medical resources. Secondly, it is of importance to strengthen the construction of multi-level medical institutions in response to public health emergencies and occupation of medical resources, based on our finding that patients in areas with higher density of health services had shorter hospital stays and hospital level was weakly related to the length of stay.

## Data Availability

The data that support our study are available from Sichuan CDC but restrictions apply to the availability of these data.

## Acknowledgements

We thank all staff in CDCs at all levels for their contributions to fight against novel coronavirus epidemic.

## Authors’ contributions

ZW, JJ, BZ and XW conceived the study design and analysis plan. ZW, YL, RL, YZ performed the statistical analysis and wrote the first draft. XC, LZ, QL, YZ, XX, TD, JZ, LZ, JH, XW, YD reviewed and revised the manuscript. All authors provided comments and suggestions and approved the final manuscript.

## Funding

This study was supported by the Grants from Science and Technology Bureau of Sichuan province (COVID-19 science and technology emergency project, No. 2020YFS0015) and Sichuan Provincial Leading Group Office for Talent Work (Sichuan provincial “Tianfu ten thousand talents plan” fund in 2018).

## Ethics approval and consent to participate

The study was approved by Ethics Committee of Sichuan CDC (SCCDCIRB-2020-006).

## Consent for publication

Not Applicable.

## Declaration of interests

All authors declare that they have no competing interests.

## Notes

### Competing Interest Statement

The authors have declared no competing interest.

